# Association Between Artificial Intelligence-Derived Tumor Volume and Oncologic Outcomes for Localized Prostate Cancer Treated with Radiation Therapy

**DOI:** 10.1101/2023.04.16.23288642

**Authors:** David D Yang, Leslie K Lee, James MG Tsui, Jonathan E Leeman, Katie N Lee, Heather M McClure, Atchar Sudhyadhom, Christian V Guthier, Kent W Mouw, Neil E Martin, Peter F Orio, Paul L Nguyen, Anthony V D’Amico, Martin T King

## Abstract

**Background:** Although clinical features of multi-parametric magnetic resonance imaging (mpMRI) have been associated with biochemical recurrence in localized prostate cancer, such features are subject to inter-observer variability.

**Objective:** To evaluate whether the volume of the dominant intraprostatic lesion (DIL), as provided by a deep learning segmentation algorithm, could provide prognostic information for patients treated with definitive radiation therapy (RT).

**Design, Setting, and Participants:** Retrospective study of 438 patients with localized prostate cancer who underwent an endorectal coil, high B-value, 3-Tesla mpMRI and were treated with RT between 2010 and 2017.

**Intervention:** RT.

**Outcome Measurements and Statistical Analysis:** Biochemical recurrence and metastasis risk, assessed with a cause-specific Cox regression and time-dependent receiver operating characteristic analysis.

**Results and Limitations:** The artificial intelligence (AI) model identified DILs with an area under the receiver operating characteristic curve (AUROC) of 0.827 at the patient level. For the 233 patients with available PI-RADS scores, with a median follow-up of 5.6 years, AI-defined DIL volume was significantly associated with biochemical failure (adjusted hazard ratio 1.54, 95% confidence interval 1.09-2.17, p=0.014) after adjustment for PI-RADS score. Among all 438 patients with a median follow-up of 6.9 years, the AUROC for predicting 7-year biochemical failure for AI volume (0.790) was similar to that for an expanded National Comprehensive Cancer Network (NCCN+) category (p=0.17). The AUROC for predicting 7-year metastasis for AI volume trended towards being higher compared to NCCN+ categories (0.854 vs 0.769, p=0.06).

**Conclusions:** A deep learning algorithm could identify the DIL with good performance. AI-defined DIL volume may be able to provide prognostic information independent of the NCCN+ risk group or other radiologic factors for patients with localized prostate cancer treated with RT.

## Introduction

Multi-parametric magnetic resonance imaging (mpMRI) has revolutionized the management of localized prostate cancer. mpMRI-based biopsy strategies have been shown to improve the detection of clinically significant disease while reducing the detection of clinically insignificant disease^1–4^. Furthermore, the characteristics of dominant intraprostatic lesions (DILs) on mpMRI, including PI-RADS scores^5^, radiologic T-stage^6,7^, and lesion size^8–11^, have been shown to be prognostic. Recently, a nomogram based on clinical and radiologic parameters was found to be highly prognostic for early biochemical recurrence after radical prostatectomy^12^.

However, the assignment of various clinical features on mpMRI is subject to significant inter-observer variability. For example, multiple grading systems^13–15^ exist for assigning extracapsular extension (EPE), each with differing sensitivities for the detection of histologic EPE extent^16^. Multi-reader studies have reported moderate inter-observer variability in the reporting of Prostate Imaging Reporting and Data Systems (PI-RADS) v2.0 scores^17–19^. A recent study demonstrated that the positive predictive value of PI-RADS scores for detecting high-grade prostate cancer was low (35%) and variable across 26 centers^20^. The varied performance of mpMRI is likely multifactorial and could be partially attributed to technical and interpretative factors, as well as benign conditions that mimic malignancy^21^.

Given these limitations, there has been considerable interest in the development of artificial intelligence (AI) algorithms for supporting the radiologist’s workflow. Although the performance of AI algorithms has not been shown to match that of radiologists for the detection of clinically significant prostate cancer^22^, recent work suggests that deep learning algorithms could be used as an adjunct for assisting radiologists^23^. However, less is known about the prognostic information provided by deep learning algorithms, particularly in comparison to current staging systems. The purpose of this study is to evaluate whether the mpMRI DIL volume, as determined by a deep learning segmentation algorithm^24^, could provide prognostic information for patients with localized prostate cancer treated with definitive radiation therapy (RT).

## Methods

### Clinical datasets

We conducted a retrospective study of 438 patients with cT1cN0M0 prostate cancer who underwent an endorectal coil, high B-value, 3-Tesla mpMRI and were treated with definitive prostate RT, both at our institution, between 2010 and 2017 (Table S1). mpMRI were obtained with either General Electric (GE) Signa HDxt (n=237) or DISCOVERY MR750w (n=201) (Table S2). Apparent diffusion coefficient (ADC)/diffusion-weighted imaging (DWI) were obtained at the same resolution (0.70 × 0.70 × 3-4 mm) as the T2-weighted images. All research was conducted with approval from the institutional review board.

We captured baseline clinical characteristics and treatment characteristics (Table 1). We stratified patients into an expanded National Cancer Center Network (NCCN+) classification, based on the 4-tiered NCCN stratification (low, favorable intermediate-risk, unfavorable intermediate-risk, high-risk)^25^, with the addition of a very high-risk category per eligibility criteria from recent STAMPEDE trials (≥2 of the following: cT3-T4, Gleason score 8-10, or PSA ≥40 ng/mL)^26^. We captured radiologic staging parameters, including PI-RADS v2.0 scores^27^, which were assessed by a subspecialty abdominal radiologist (L.K.L.) for 83 patients who were scanned in 2010-2013 and available for all patients after mid-2015, as well as radiologic T-stage (T1: no visible tumor on mpMRI, T2: organ-confined, T3a: EPE, T3b: seminal vesicle invasion [SVI]) based on mpMRI. Lastly, we captured outcomes including biochemical failure (nadir+2 ng/mL), development of metastasis (non-regional nodal or distant disease on bone scan, computed tomography, or positron emission tomography), and survival.

**Table 1:** Baseline clinical, radiologic, and treatment factors. SV: seminal vesicles. EBRT: external beam radiation therapy. Radiologic T stage and hemorrhage data were not available for the TestNoPIRADS cohort.

| Characteristics | TrainPIRADS (n=150) | TestPIRADS (n=83) | TestNoPIRADS (n= 205) |
| --- | --- | --- | --- |
| Age<br>Median (IQR) | 69.5 (64.0, 73.0) | 68.0 (65.0, 73.0) | 66 (61.0, 72.0) |
| Gleason grade group |  |  |  |
| 1 | 18 (12.0%) | 17 (20.5%) | 90 (43.9%) |
| 2 | 62 (41.3%) | 32 (38.6%) | 70 (34.2%) |
| 3 | 30 (20.0%) | 11 (13.3%) | 23 (11.2%) |
| 4 | 22 (14.7%) | 9 (10.8%) | 9 (4.4%) |
| 5 | 18 (12.0%) | 14 (16.9%) | 13 (6.3%) |
| Clinical T-stage |  |  |  |
| T1-T2a | 122 (81.3%) | 69 (83.1%) | 182 (88.8%) |
| T2b-T2c | 12 (8.0%) | 10 (12.1%) | 17 (8.3%) |
| T3a | 8 (5.3%) | 3 (3.6%) | 5 (2.4%) |
| T3b | 8 (5.3%) | 1 (1.2%) | 1 (0.5%) |
| PSA |  |  |  |
| 0-9.9 | 107 (71.3%) | 63 (75.9%) | 179 (87.3%) |
| 10.0-19.9 | 27 (18.0%) | 12 (14.5%) | 19 (9.3%) |
| 20.0+ | 16 (10.7%) | 8 (9.6%) | 7 (3.4%) |
| PPC |  |  |  |
| 0-49% | 80 (53.3%) | 49 (59.0%) | 161 (78.5%) |
| 50-100% | 65 (43.3%) | 33 (39.8%) | 41 (20.0%) |
| Missing | 5 (3.3%) | 1 (1.2%) | 3 (1.5%) |
| NCCN+ category |  |  |  |
| Low | 13 (8.7%) | 14 (16.9%) | 80 (39.0%) |
| FIR | 34 (22.7%) | 14 (16.9%) | 57 (27.8%) |
| UIR | 52 (34.7%) | 24 (28.9%) | 38 (18.5%) |
| High | 37 (24.7%) | 28 (33.7%) | 26 (12.7%) |
| Very High | 14 (9.3%) | 3 (3.61) | 4 (2.0%) |
| Model |  |  |  |
| Signa HDxt | 52 (34.7%) | 31 (37.3%) | 154 (75.1%) |
| DISCOVERY |  |  |  |
| MR750w | 98 (65.3%) | 52 (62.7%) | 51 (24.9%) |
| PIRADS |  |  |  |
| 0-2 | 6 (4.0%) | 10 (12.1%) | NA |
| 3 | 15 (10.0%) | 8 (9.6%) | NA |
| 4 | 48 (32.0%) | 24 (28.9%) | NA |
| 5 | 81 (54.0%) | 41 (49.4%) | NA |
| Radiologic T-stage |  |  |  |
| T1 | 6 (4.00%) | 10 (12.05%) | N/A |
| T2 | 83 (55.33%) | 49 (59.04%) | N/A |
| T3a | 45 (30.00%) | 20 (24.10%) | N/A |
| T3b | 16 (10.67%) | 4 (4.82%) | N/A |
| Hemorrhage |  |  |  |
| Yes | 56 (37.33%) | 28 (33.73%) | N/A |

Table 1: Baseline clinical, radiologic, and treatment factors.
| No | 94 (62.67%) | 55 (66.27%) | N/A |
| --- | --- | --- | --- |
| Time between biopsy and MRI (days) |  |  |  |
| Patients with available data | 148 | 82 | 204 |
| Median (IQR) | 46.5 (24.0, 72.3) | 48.5 (22.5, 101.3) | 36.0 (19.0, 59.3) |
| Time between MRI and Treatment (days) |  |  |  |
| Median (IQR) | 23.0 (7.3, 50.3) | 44.0 (15.5, 66.0) | 54.0 (27.0, 83.0) |
| Radiation Therapy |  |  |  |
| EBRT | 110 (73.3%) | 54 (65.1%) | 83 (40.5%) |
| Brachytherapy | 21 (14.00%) | 19 (22.9%) | 117 (57.1%) |
| Combination | 19 (12.7%) | 10 (12.0%) | 5 (2.4%) |
| Radiation Nodal Coverage |  |  |  |
| Prostate/SV | 132 (88.0%) | 71 (85.5%) | 199 (97.1%) |
| Pelvic Nodes | 18 (12.0%) | 12 (14.5%) | 6 (2.9%) |
| Prostate EBRT Dose (cGy) |  |  |  |
| 7020 | 8 (5.33%) | 5 (6.02%) | 8 (3.90%) |
| 7560-7920 | 100 (66.67%) | 49 (59.04%) | 75 (36.59%) |
| 6000 | 2 (1.33%) | 0 (0.00%) | 0 (0.00%) |
| ADT Duration (mo) |  |  |  |
| 0-2.9 | 36 (24.0%) | 28 (33.7%) | 126 (61.4%) |
| 3.0-6.0 | 71 (47.3%) | 31 (37.4%) | 53 (25.9%) |
| 6.1-18.0 | 7 (4.7%) | 4 (4.8%) | 2 (1.0%) |
| >18.0 | 36 (24.0%) | 20 (24.1%) | 24 (11.7%) |
| Systemic therapy Intensification |  |  |  |
| None | 138 (92.0%) | 77 (92.8%) | 198 (96.5%) |
| Docetaxel | 2 (1.3%) | 1 (1.2%) | 4 (2.0%) |
| Enzalutamide | 10 (6.7%) | 5 (6.0%) | 3 (1.5%) |
| Year of Treatment Initiation |  |  |  |
| 2010-2014 | 52 (34.7%) | 31 (37.3%) | 187 (91.2%) |
| 2015-2017 | 98 (65.3%) | 52 (62.7%) | 18 (8.8%) |

We divided patients into 3 cohorts. Of the 233 patients with available PI-RADS scores, we randomly allocated them into training (TrainPIRADS) and testing (TestPIRADS) sets. The remaining 205 patients were allocated into a second testing (TestNoPIRADS) set. For the TrainPIRADS and TestPIRADS sets, a radiation oncologist contoured the prostate transitional zone (TZ), peripheral zone (PZ), and DIL based on the presence of PI-RADS 3-5 scores. For the TestNOPIRADS set, the radiation oncologist contoured only the DIL based on radiology reports and biopsy data.

### nnUNet algorithm

We utilized the publicly available nnUNet^24^ deep learning algorithm for automated delineations of the TZ, PZ, and DIL. First, T2, ADC, and DWI images were downloaded onto a research workstation. Images were anonymized and cropped into smaller arrays (128 × 128 × 15 or 20 pixels). T2-weighted images were registered onto the ADC images, utilizing the best performing registration among 4 algorithms (rigid, rigid + B-spline, affine, affine + B-spline) of prostate segmentation distance maps^28^. The median registration Dice coefficient was 0.92 (interquartile range [IQR] 0.90-0.94).

We trained a 3D full-resolution nnUNet model utilizing 5-fold cross-validation on the TrainPIRADS set. We then applied the selected configuration onto the TestPIRADS and TestNoPIRADS sets. With the utilization of 5-fold cross-validation on the training set and application of the model on the 2 testing sets, we generated TZ, PZ, and DIL segmentations for all 438 patients.

### Assessment of AI DIL volume performance

We reported the Dice coefficients for the TZ and PZ, as noted by nnUNet. For the DIL, we analyzed the detection performance (patient-level area under the receiver operator characteristic curve [AUROC] and lesion-level specificity at 0.5 false positive per image) utilizing PICAI_eval^29^. To assess whether false positive (FP) or false negative (FN) lesions were smaller and less conspicuous than true positives (TP), we classified AI lesions as TP, FP, or FN, based on whether a reference DIL was present and correctly identified (TP: Dice coefficient ≥10%^29^), identified but not present (FP), or present but not identified or with insufficient Dice coefficient overlap (<10%) (FN). We analyzed lesion-level DIL volumes and contrast ratios (ratio of ADC intensities within the DIL versus a surrounding 2mm ring) based on classification status. Finally, we reported the similarity metrics (Dice coefficient) for TP lesions. All processing was performed using Python version 3.8.12 with the SimpleITK toolkit.

### Comparison of AI and reference DIL volumes for sextant subset

For the 303 patients with sextant biopsies among all 3 cohorts, we compared the ability of the AI and reference DIL contours to detect clinically significant (Gleason grade group ≥2) disease within each sextant. We compared area under the curve (AUC) values utilizing ROC analysis (R package pROC). AI and reference DIL contours were allocated according to sextants based on the reference prostate contour.

### Comparison of DIL volume with radiologic staging for PI-RADS subset

For the 233 patients (TrainPIRADS and TestPIRADS) with available PI-RADS scores among all 3 cohorts, we conducted cause-specific Cox regression models (R package coxph) for predicting biochemical failure with log-transformed AI DIL volume in cm^3^ (V_AI_), clinical T-stage, radiologic T-stage, PI-RADS scores (0-4 vs 5^30^), age, whether patients received standard of care (SOC, i.e. ≥75.6 Gy if receiving external beam RT and androgen deprivation therapy [ADT] duration of 6 months for unfavorable intermediate-risk disease and ≥18 months for high/very high-risk disease), year of treatment, and scanner model (Signa HDxt vs DISCOVERY MR750w) as independent variables. An additional Cox regression model for metastasis was created using the above independent variables with the addition of salvage ADT use as a time-dependent variable.

### Comparison of DIL volume with NCCN+ risk category for entire cohort

For the entire cohort, we created ROC curves for NCCN+, reference DIL volume (V_REF_), and V_AI_ for predicting 7-year biochemical failure and metastasis, utilizing the timeROC package. We also created cause-specific Cox regression models (R package coxph) for predicting risk of biochemical failure, local failure, and metastasis. Independent variables included log-transformed V_AI_, NCCN+ classification, age, receipt of SOC, year of treatment, scanner model, and for local failure and metastasis models, salvage ADT use as a time-dependent variable. Finally, we estimated cumulative incidences of biochemical failure, local failure, and metastasis, based on partitions of V_AI_ intervals, utilizing the R package prodlim. All statistical analysis was conducted with R version 4.1.2.

## Results

### Assessment of AI DIL performance

For the TestPIRADS cohort, median Dice coefficients for the TZ and PZ were 0.89 and 0.84, respectively (Table S3). The nnUNet exhibited a patient-level AUROC of 0.827 and a lesion-level sensitivity of 72.7% at a false positive rate of 0.5 case per image (Table S4). For patient-level analysis (Table S5), there was no significant difference between V_AI_ (median 1.02cm^3^, IQR 0.40-2.34) and V_REF_ (median 1.31cm^3^, IQR 0.37-2.61, p=0.66), with Pearson’s correlation coefficient of 0.79. For lesion-level analysis (Table S6), the total volume of AI TP lesions (median 1.06cm^3^, IQR 0.47-1.85) was significantly greater than for FP (median 0.25cm^3^, IQR 0.09-0.58; p<0.001) and FN (median 0.37cm^3^, IQR 0.21-0.88, p=0.007) lesions. The TP lesions were also more conspicuous (median contrast ratio 0.72, IQR 0.68-0.76) than FP (median 0.83, IQR 0.79-0.86; p<0.001) and FN (median 0.81, IQR 0.79-0.88; p<0.001) lesions. The median Dice coefficient for TP lesions was 0.74. Examples of TP, FP, and FN lesions are shown in Fig. S1.

### Comparison of AI and reference DIL contours for sextant subset

For the subset of 303 patients with sextant biopsies among all 3 cohorts, we did not detect a difference in AUROC values associated with AI versus reference contours for any sextant (Table S7).

### Comparison of DIL volume with radiologic staging for PI-RADS subset

For the 233 patients with PI-RADS scores (TrainPIRADS and TestPIRADS cohorts), with median follow-up of 5.6 years, there were 28 biochemical failures. AI algorithm detected 41.2%, 68.3%, and 87.1% of PI-RADS3, 4, and 5 lesions, respectively (Table S8). PIRADS 5 vs 0-4 (HR 5.90, 95% confidence interval [CI] 2.05-17.03, p=0.001) and V_AI_ (HR 1.89 per log cm^3^ increase, 95% CI 1.41-2.53, p<0.001) were significantly associated with biochemical failure on univariable analysis (Table 2). However, only V_AI_ retained significance on multivariable analysis (adjusted HR 1.54, 95% CI 1.09-2.17; p=0.014). The association between V_AI_ and metastasis was similarly significant (adjusted HR 1.71, 95% CI 1.04-2.80, p=0.04; Table S9).

**Table 2:**
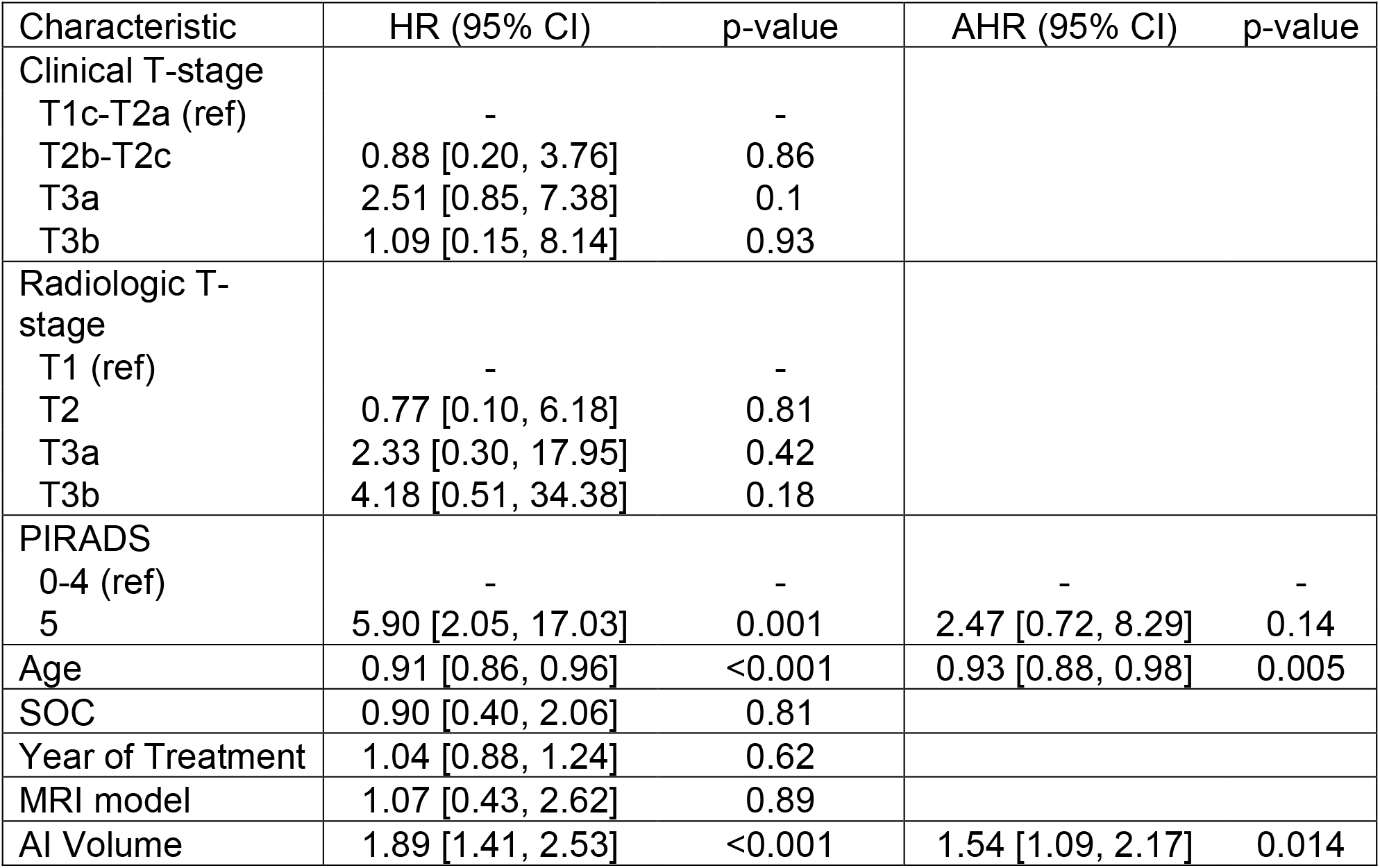
Cox regression model for biochemical failure for the subset of 233 patients with available PI-RADS scores. CI: confidence interval. HR: hazard ratio. AHR: adjusted hazard ratio. SOC: standard of care treatment. Ref: reference.

| Characteristic | HR (95% CI) | p-value | AHR (95% CI) | p-value |
| --- | --- | --- | --- | --- |
| Clinical T-stage |  |  |  |  |
| T1c-T2a (ref) | - | - |  |  |
| T2b-T2c | 0.88 [0.20, 3.76] | 0.86 |  |  |
| T3a | 2.51 [0.85, 7.38] | 0.1 |  |  |
| T3b | 1.09 [0.15, 8.14] | 0.93 |  |  |
| Radiologic T-stage |  |  |  |  |
| T1 (ref) | - | - |  |  |
| T2 | 0.77 [0.10, 6.18] | 0.81 |  |  |
| T3a | 2.33 [0.30, 17.95] | 0.42 |  |  |
| T3b | 4.18 [0.51, 34.38] | 0.18 |  |  |
| PIRADS |  |  |  |  |
| 0-4 (ref) | - | - | - | - |
| 5 | 5.90 [2.05, 17.03] | 0.001 | 2.47 [0.72, 8.29] | 0.14 |
| Age | 0.91 [0.86, 0.96] | <0.001 | 0.93 [0.88, 0.98] | 0.005 |
| SOC | 0.90 [0.40, 2.06] | 0.81 |  |  |
| Year of Treatment | 1.04 [0.88, 1.24] | 0.62 |  |  |
| MRI model | 1.07 [0.43, 2.62] | 0.89 |  |  |
| AI Volume | 1.89 [1.41, 2.53] | <0.001 | 1.54 [1.09, 2.17] | 0.014 |

### Comparison of DIL volume with NCCN+ risk category for the entire cohort

Among all 438 patients with a median follow-up of 6.9 years, there were 49 biochemical failures, 14 local failures, and 22 metastases. The AUROC for 7-year biochemical failure for V_AI_ (0.790) was similar to that for V_REF_ (0.779; p=0.42) and NCCN+ category (0.740; p=0.17). The sensitivity and specificity for 7-year biochemical failure at DIL volumes >0.5 mL were 90.5% and 45.8%, respectively, and at DIL volumes >2.0 mL, were 57.7% and 84.4%, respectively. The AUROC for predicting 7-year metastasis for V_AI_ (0.854) was similar to that for V_REF_ (0.817; p=0.15) but trended towards being higher compared to NCCN+ category (0.769; p=0.06). The sensitivity and specificity for predicting 7-year metastasis at DIL volumes >0.5 mL were 94.3% and 44.1%, respectively, and at DIL volumes >2.0 mL were 68.1% and 82.9%, respectively (Fig. 1).

**Fig 1:**
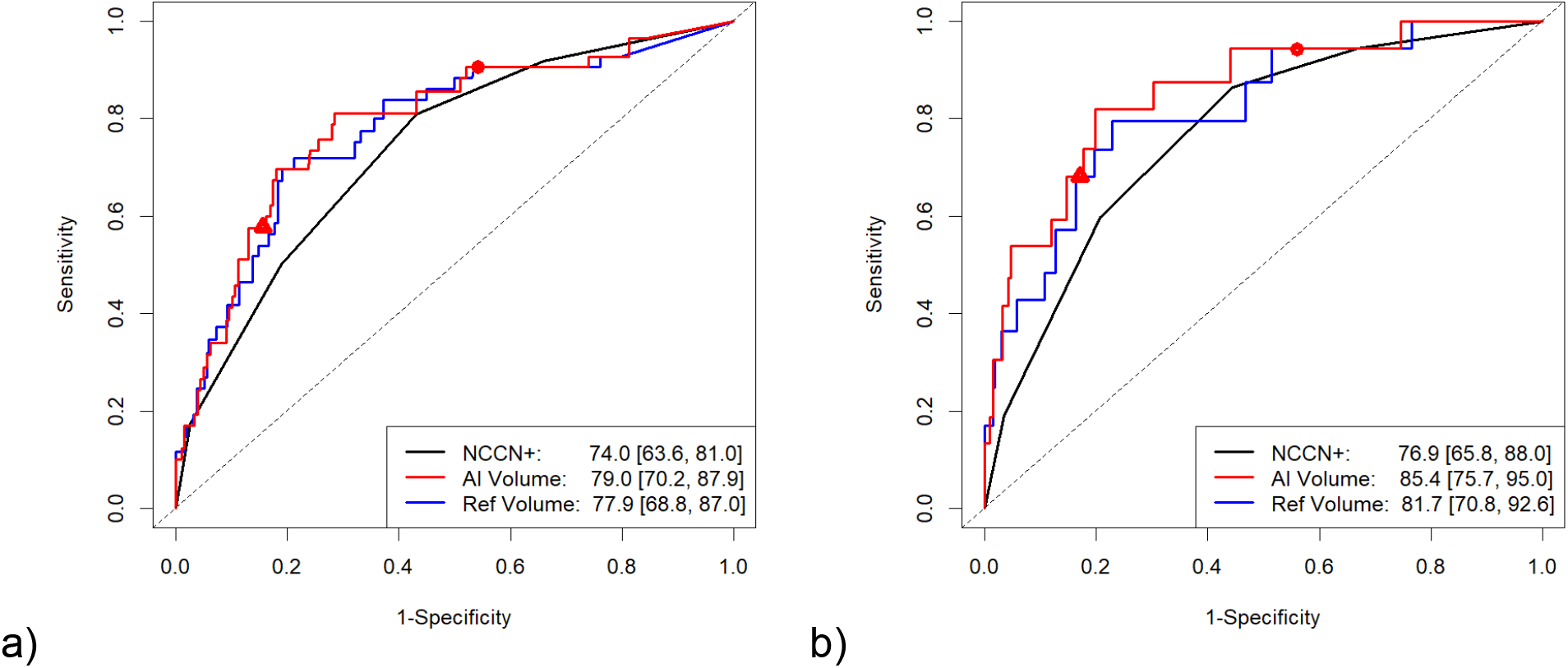
Receiver operating characteristics curves for 7-year biochemical failure (a) and metastasis (b), comparing AI DIL volume (red) against reference DIL volume (blue) and NCCN+ stratification (black). Circle and triangle are points on the AI volume receiver operating characteristics curve corresponding to thresholds of 0.5 cc and 2.0 cc, respectively.

V_AI_ was significant on both univariable and multivariable analyses for predicting biochemical failure (Table 3; adjusted HR 1.61, 95% CI 1.23-2.12, p<0.001). For metastasis, only V_AI_ was significant on univariable and multivariable analysis (Table 4; adjusted HR, 1.71, 95% CI 1.08-2.70, p=0.02). V_AI_ was also associated with local recurrence risk on univariable analysis (HR 1.41, 95% CI 1.00-1.98, p=0.05; Table S10).

**Table 3:**
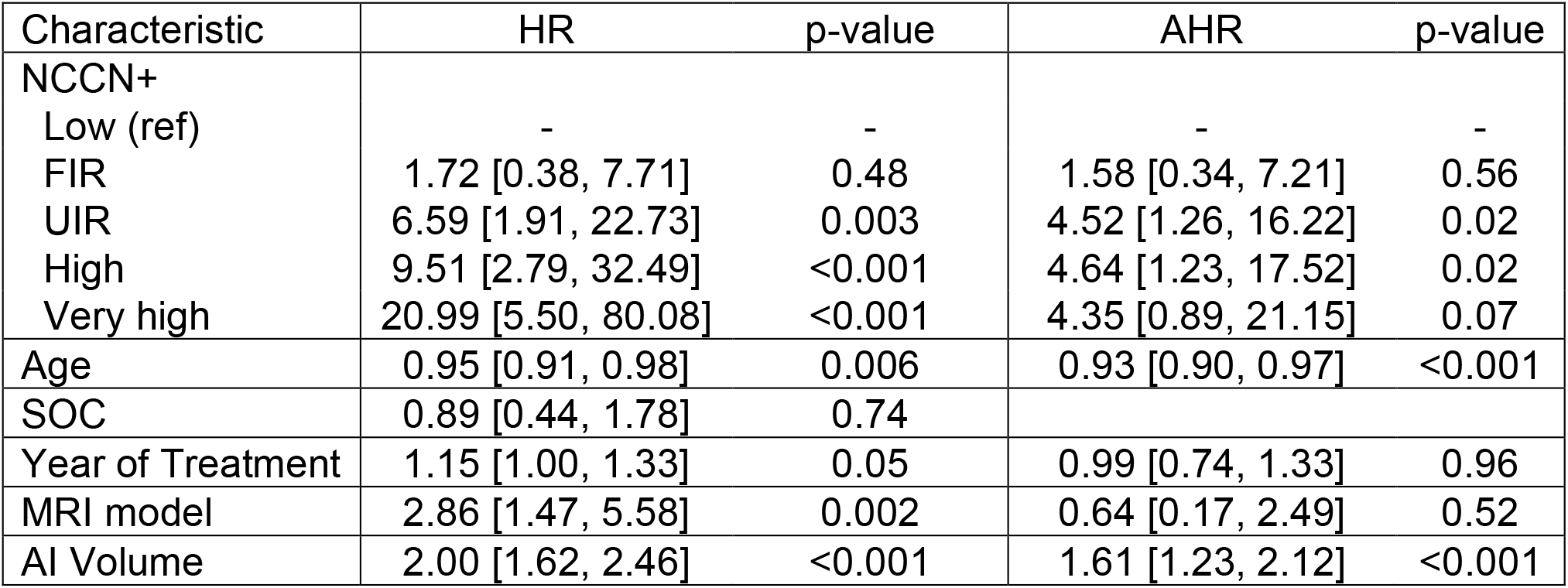
Cox regression model for biochemical failure for all patients. CI: confidence interval. HR: hazard ratio. AHR: adjusted hazard ratio. SOC: standard of care treatment. Ref: reference. FIR: favorable intermediate-risk. UIR: unfavorable intermediate-risk.

**Table 4:** Cox regression model for metastasis for all patients. CI: confidence interval. HR: hazard ratio. AHR: adjusted hazard ratio. SOC: standard of care treatment. Ref: reference. FIR: favorable intermediate-risk. UIR: unfavorable intermediate-risk.

| Characteristic | HR (95% CI) | p-value | AHR (95% CI) | p-value |
| --- | --- | --- | --- | --- |
| NCCN+ |  |  |  |  |
| Low/FIR (ref) | - | - | - | - |
| UIR | 3.07 [0.68, 13.76] | 0.14 | 1.33 [0.27, 6.48] | 0.73 |
| High | 10.63 [2.95, 38.29] | <0.001 | 3.18 [0.75, 13.60] | 0.12 |
| Very high | 17.84 [3.94, 80.71] | <0.001 | 1.47 [0.21, 10.27] | 0.7 |
| Age | 0.96 [0.90, 1.02] | 0.016 |  |  |
| SOC | 1.04 [0.35, 3.08] | 0.94 |  |  |
| Year of Treatment | 1.23 [0.97, 1.55] | 0.08 |  |  |
| MRI model | 5.36 [1.70, 16.86] | 0.004 | 0.53 [0.17, 1.66] | 0.27 |
| Salvage ADT | 29.53 [11.19, 77.92] | <0.001 | 12.58 [4.51, 31.11] | <0.001 |
| AI Volume | 2.29 [1.67, 3.15] | <0.001 | 1.71 [1.08, 2.70] | 0.02 |

Fig. 2 shows cumulative incidences of biochemical failure, local failure, and metastasis for V_AI_ of 0-0.4, 0.5-1.9, and ≥2.0cm^3^. Respective incidences of 7-year biochemical failure were 2.7%, 9.2%, and 23.7%. Respective incidences of 7-year local failure were 1.0%, 5.8%, and 5.3%. Respective incidences of 7-year metastasis were 0.7%, 2.9%, and 11.8%. Table S11 depicts the association of DIL volume ranges with NCCN+ and radiologic factors. Cumulative incidence curves for NCCN+ categories are shown in Fig. S2.

**Fig 2:**
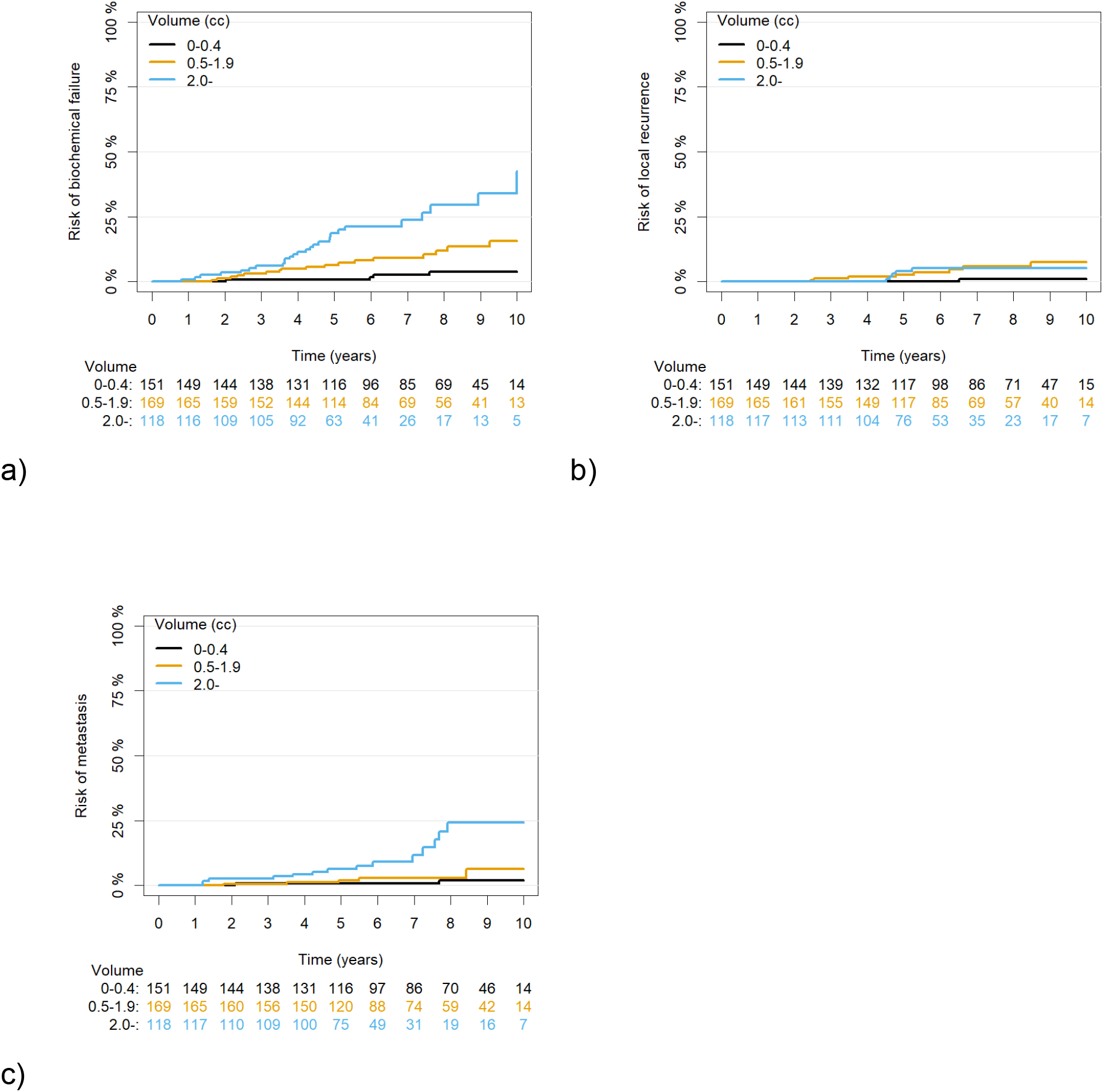
Cumulative incidence curves of DIL volume intervals (0-0.4, 0.5-1.9, 2.0-cc) with (a) biochemical failure, (b) local failure, and (c) metastasis.

## Discussion

Our study demonstrates that the prostate and DIL can be accurately delineated on mpMRI using nnUNet,^24^ an out-of-the-box self-configuring deep learning segmentation algorithm. Our findings suggest that AI-determined DIL volume provides prognostic information comparable to both NCCN+ risk category and human-generated DIL volume for estimating biochemical recurrence and metastasis risk for localized prostate cancer treated with RT. Furthermore, for the subset of patients with PI-RADS scores, V_AI_ was more strongly associated with biochemical failure than other mpMRI parameters, including radiologic staging and PI-RADS score.

Though other studies have reported on the prognostic significance of tumor size for biochemical recurrence^8–11^, ours is among the first to show that DIL volume can be reliably obtained with an AI algorithm. This is clinically meaningful because DIL volume determination can be time-consuming and is subject to significant inter-observer variability^31^. The AI algorithm, on the other hand, was able to generate the DIL volume in an efficient, automated, and standardized manner. Additionally, this work may be clinically impactful as it suggests that V_AI_ could provide important prognostic information even in the absence of a biopsy. With further validation, V_AI_ could have the potential to provide unique prognostic information in addition to that provided with current clinical and radiologic staging systems.

V_AI_ shows exceptional promise as a potential prognostic factor as it is a single, well-defined entity which may be generated in a systematic manner from mpMRI. Unlike NCCN risk categories^25^, V_AI_ does not require the presence of a systematic biopsy, which may be important as MR-only biopsies are increasingly utilized^4^. Unlike radiomic approaches, V_AI_ does not require the selection and regression of multiple features, which may have limited generalizability due to inter- and intra-scanner reproducibility^32^. Unlike genomic classifiers^33^ or digital pathology approaches^34^, no specific retrieval or handling of clinical specimens are required. Rather, prognostic information could be obtained directly from the mpMRI.

Additionally, this study provides support for extreme dose-escalation to large DILs, either through external beam RT^35^ or brachytherapy^36,37^ techniques. We found that V_AI_ to be associated with local recurrence risk. Given emerging evidence which strongly suggests that local failures seed distant metastases^38^, extreme dose-escalation in appropriately selected patients with large DILs may be clinically impactful.

Our study showed that FP and FN lesions were smaller and less conspicuous than TP lesions. Our AI algorithm may have retained prognostic significance, despite missing such lesions, because of the weaker association between smaller lesions^8–11^ and less conspicuous ADC values^39^ with biochemical recurrence.

Key strengths of this study include the utilization of a well-annotated contemporary patient cohort treated with modern RT approaches. Furthermore, this study shows that mpMRI obtained after prior biopsy and with an endorectal coin contains important prognostic information that can be extracted by deep learning approaches. At our institution and likely many others, most mpMRIs acquired before 2018 had a pre-existing biopsy^1^.

It should be noted that the algorithm developed within this study may not be generalizable to MRIs obtained using different scanning parameters or without endorectal coils. External validation would be necessary to establish generalizability. It is our hope that deep learning algorithms with similar levels of performance may be built using nnUNet (or other deep learning segmentation approaches) for MRIs obtained under different conditions. Additionally, the AI algorithm missed significant disease in a subset of cases, including 12.9% of PI-RADS 5 lesions. As a result, AI DIL volume should not be used as a standalone entity.

Further research involves extending this work to multi-institutional datasets involving other scanning configurations and treatment types (e.g. surgery). We are very encouraged by the excellent performance of deep learning algorithms across multi-institution datasets^21,28^ and look forward to the results of the PICAI-Eval challenge^29^.

Only with additional information can we better understand the full potential of deep learning AI algorithms for providing potentially valuable prognostic information for patients.

## Data Availability

All data produced in the present study are available upon reasonable request to the authors.

