## Supplemental Figures and Tables for "Association Between Artificial Intelligence-Derived Tumor Volume and Oncologic Outcomes for Localized Prostate Cancer Treated with Radiation Therapy"

Table S1: Formation of the patient cohort.

| | | | |
|---|---|---|---|
| 3161 | patients who underwent prostate RT at BWH between 1/09-1/18 | | |
| 867 | Pre-RT mpMRI | | |
| 764 | Pre-RT mpMRI at BWH | | |
| 704 | Pre-RT mpMRI at BWH with GE scanner (high-resolution diffusion weighted images (DWI)) | | |
| 497 | Pre-RT high B-value (>= 1400) DWI mpMRI with GE scanner | | |
| | 59 | Excluded for the following reasons | |
| | | 1 | 1.5T interventional MRI |
| | | 10 | Hip artifacts |
| | | 1 | Large hematoma |
| | | 4 | MRI > 1 month on ADT |
| | | 4 | MRI > 1 year old |
| | | 4 | N1 disease |
| | | 11 | No endorectal coil |
| | | 1 | Partial prostatectomy |
| | | 1 | Penile implant |
| | | 11 | Prior ADT |
| | | 1 | Prior microwave ablation |
| | | 1 | Prior sarcoid |
| | | 8 | Severe DWI artifacts |
| | | 1 | Wide FOV (pelvis) |
| 438 | Final cohort | | |

Table S2: Scanning acquisition parameters. All diffusion-weighted imaging (DWI) scans were obtained with a B-value ≥ 1000 s/mm$^2$ with an endorectal coil.

| Characteristic | Signa HDxt (N = 237) | DISCOVERY MR750w (N = 201) |
|---|---|---|
| Acquisition | | |
| Echo Time | 2,500.00 (2,500.00, 2,500.00) | 7,550.00 (7,300.00, 7,550.00) |
| Repetition Time | 81.20 (80.60, 81.40) | 78.80 (77.90, 79.20) |
| Field Strength | 3.00 (3.00, 3.00) | 3.00 (3.00, 3.00) |
| Image | | |
| Pixel spacing | 0.70 (0.70, 0.70) | 0.70 (0.70, 0.70) |
| Slice spacing | 3.00 (3.00, 3.00) | 4.00 (4.00, 4.00) |
| Recon Diameter | 180.00 (180.00, 180.00) | 179.99 (179.99, 179.99) |

Table S3: Performance of AI algorithm for segmenting the TZ and PZ.

| | TrainPIRADS (N = 150) | TestPIRADS (N = 83) |
|---|---|---|
| TZ Dice Median (IQR) | 0.88 (0.83, 0.92) | 0.89 (0.84, 0.92) |
| PZ Dice Median (IQR) | 0.83 (0.75, 0.88) | 0.84 (0.79, 0.87) |

Table S4: nnUNet performance.

| Performance | TrainPIRADS (N = 150) | TestPIRADS (N = 83) | TestNOPIRADS (N = 205) |
|---|---|---|---|
| Patient-level AUROC | 0.906 | 0.827 | 0.785 |
| Sensitivity at 0.5 FP | 71.8% | 72.7% | 65.8% |
| Average Precision | 67.8% | 67.0% | 56.8% |

Table S5: Patient-specific volume characteristics.

| Patient-level characteristics | TrainPIRADS (N = 150) | TestPIRADS (N = 83) | TestNOPIRADS (N = 205) |
|---|---|---|---|
| Classification | | | |
| Num TP | 128 (85.33%) | 62 (74.70%) | 117 (57.07%) |
| Num FP | 5 (3.33%) | 9 (10.84%) | 36 (17.56%) |
| Num FN | 16 (10.67%) | 11 (13.25%) | 32 (15.61%) |
| Num TN | 1 (0.67%) | 1 (1.20%) | 20 (9.76%) |
| Num Ref DIL | | | |
| 0 | 6 (4.00%) | 10 (12.05%) | 56 (27.32%) |
| 1 | 97 (64.67%) | 49 (59.04%) | 118 (57.56%) |
| 2 | 36 (24.00%) | 20 (24.10%) | 19 (9.27%) |
| ≥3 | 11 (7.33%) | 4 (4.82%) | 12 (5.85%) |
| Num Test DIL | | | |
| 0 | 5 (3.33%) | 6 (7.23%) | 36 (17.56%) |
| 1 | 89 (59.33%) | 45 (54.22%) | 120 (58.54%) |
| 2 | 44 (29.33%) | 25 (30.12%) | 41 (20.00%) |
| ≥3 | 12 (8.00%) | 7 (8.43%) | 8 (3.90%) |
| Total Volume ($cm^3$) | | | |
| Reference | 1.84 (0.78, 4.81) | 1.31 (0.37, 2.61) | 0.28 (0.00, 0.97) |
| Test | 1.50 (0.71, 3.51) | 1.02 (0.40, 2.34) | 0.52 (0.10, 1.38) |

Table S6: Lesion-specific volume characteristics.

| Lesion-level characteristics | TrainPIRADS (N = 271) | TestPIRADS (N = 145) | TestNOPIRADS (N = 299) |
|---|---|---|---|
| Classification | | | |
| Num TP | 145 (53.1%) | 73 (50.3%) | 129 (43.1%) |
| Num FP | 69 (25.5%) | 44 (30.3%) | 103 (34.5%) |
| Num FN | 57 (21.0%) | 28 (19.3%) | 67 (22.4%) |
| Volume | | | |
| All | 0.59 (0.20, 1.78) | 0.57 (0.23, 1.50) | 0.32 (0.11, 0.82) |
| TP (nonzero) | 145; 1.23 (0.58, 3.54) | 73; 1.06 (0.47, 1.85) | 129; 0.70 (0.32, 1.51) |
| FP (nonzero) | 69; 0.18 (0.08, 0.33) | 44; 0.25 (0.09, 0.58) | 103; 0.22 (0.10, 0.45) |
| FN (nonzero) | 57; 0.31 (0.19, 0.69) | 28; 0.37 (0.21, 0.88) | 67; 0.11 (0.07, 0.25) |
| Contrast | | | |
| All | 0.77 (0.72, 0.83) | 0.78 (0.71, 0.84) | 0.77 (0.71, 0.82) |
| TP (nonzero) | 145; 0.73 (0.68, 0.78) | 73; 0.72 (0.68, 0.76) | 129; 0.73 (0.69, 0.78) |
| FP (nonzero) | 69; 0.84 (0.76, 0.88) | 44; 0.83 (0.79, 0.86) | 103; 0.81 (0.77, 0.85) |
| FN (nonzero) | 57; 0.82 (0.77, 0.86) | 28; 0.81 (0.79, 0.88) | 67; 0.78 (0.74, 0.83) |
| Metrics for TP | | | |
| Dice Median (IQR) | 145; 0.77 (0.64, 0.84) | 73; 0.74 (0.58, 0.82) | 129; 0.71 (0.56, 0.79) |

Table S7: Detection of clinically significant disease by the AI versus reference contours for subset of cases 303 cases across all 3 cohorts with sextant biopsies.

| Location | AI contour AUC | Ref contour AUC | p-value |
|---|---|---|---|
| Left apex | 0.71 (0.64, 0.77) | 0.74 (0.67, 0.80) | 0.24 |
| Right apex | 0.73 (0.69, 0.80) | 0.76 (0.70, 0.83) | 0.29 |
| Left mid | 0.70 (0.64, 0.77) | 0.70 (0.64, 0.76) | 0.81 |
| Right mid | 0.77 (0.71, 0.83) | 0.78 (0.72, 0.84) | 0.75 |
| Left base | 0.68 (0.62, 0.74) | 0.65 (0.59, 0.71) | 0.39 |
| Right base | 0.68 (0.62, 0.75) | 0.72 (0.65, 0.79) | 0.14 |

Table S8: Lesion-specific characteristics of AI contours for PI-RADS 3-5 lesions.

| PI-RADS lesion characteristics | 3 (N = 51) | 4 (N = 120) | 5 (N = 132) |
|---|---|---|---|
| Lesion-Level Metric | | | |
| Num TP | 21 (41.2%) | 82 (68.3%) | 115 (87.1%) |
| Num FN | 30 (58.8%) | 38 (31.7%) | 17 (12.9%) |
| Performance | | | |
| Sensitivity | 41.2% | 68.3% | 87.1% |
| Volume | 0.28 (0.14, 0.65) | 0.51 (0.26, 0.81) | 2.87 (1.20, 4.84) |
| Contrast | 0.83 (0.78, 0.86) | 0.76 (0.71, 0.81) | 0.72 (0.67, 0.77) |
| Metrics for TP | | | |
| Dice Median (IQR) | 21; 0.47 (0.41, 0.74) | 82; 0.74 (0.60, 0.81) | 115; 0.78 (0.69, 0.84) |

Table S9: Cox regression for factors associated with time to metastasis for the subset of 233 patients with available PI-RADS scores.

CI: confidence interval. HR: hazard ratio. AHR: adjusted hazard ratio. SOC: standard of care treatment. Ref: reference.

| Characteristic | HR | p-value | AHR | P-value |
|---|---|---|---|---|
| Clinical T-stage | | | | |
| T1c-T2c (ref) | - | - | | |
| T3a-T3b | 1.99 [0.43, 9.25] | 0.38 | | |
| Radiologic T-stage | | | | |
| T1-T2 (ref) | - | - | | |
| T3a-T3b | 2.70 [0.79, 9.25] | 0.11 | | |
| PIRADS | | | | |
| 0-4 (ref) | - | - | | |
| 5 | 4.00 [0.86, 18.53] | 0.08 | | |
| Age | 0.91 [0.84, 0.99] | 0.04 | 0.94 [0.87, 1.03] | 0.018 |
| SOC | 0.66 [0.19, 2.27] | 0.51 | | |
| Year of Treatment | 1.05 [0.80, 1.39] | 0.72 | | |
| MRI model | 1.17 [0.26, 5.14] | 0.83 | | |
| Salvage ADT | 9.95 [1.88, 52.65] | 0.007 | 3.85 [0.65, 22.80] | 0.14 |
| AI Volume | 1.88 [1.17, 3.00] | 0.009 | 1.71 [1.04, 2.80] | 0.04 |

Table S10: Cox regression model for local recurrence.

CI: confidence interval. HR: hazard ratio. AHR: adjusted hazard ratio. SOC: standard of care treatment. Ref: reference. FIR: favorable intermediate-risk. UIR: unfavorable intermediate-risk.

| Characteristic | HR | p-value | AHR | p-value |
|---|---|---|---|---|
| NCCN+ | | | | |
| Low/FIR (ref) | - | - | | |
| UIR/High/Very high | 1.85 [0.62, 5.55] | 0.27 | | |
| Age | 0.92 [0.85, 0.99] | 0.02 | 0.92 [0.85, 0.99] | 0.02 |
| SOC | 0.86 [0.24, 3.08] | 0.82 | | |
| Year of Treatment | 1.07 [0.82, 1.39] | 0.64 | | |
| MRI model | 1.52 [0.49, 4.70] | 0.47 | | |
| Salvage ADT | 4.51 [1.00, 20.38] | 0.05 | 1.73 [0.31, 9.62] | 0.53 |
| AI Volume | 1.41 [1.00, 1.98] | 0.05 | 1.34 [0.95, 1.89] | 0.09 |

Table S11: Association of AI DIL volume ranges with baseline clinical and radiographic factors

| Characteristic | 0-0.4 (N = 151) | 0.5-1.9 (N = 169) | 2.0- (N = 118) | p-value |
|---|---|---|---|---|
| NCCN+ category | | | | <0.001 |
| Low | 58 (38.41%) | 45 (26.63%) | 4 (3.39%) | |
| FIR | 48 (31.79%) | 43 (25.44%) | 14 (11.86%) | |
| UIR | 31 (20.53%) | 49 (28.99%) | 34 (28.81%) | |
| High | 14 (9.27%) | 29 (17.16%) | 48 (40.68%) | |
| Very High | 0 (0.00%) | 3 (1.78%) | 18 (15.25%) | |
| Model | | | | <0.001 |
| Signa HDxt | 114 (75.50%) | 89 (52.66%) | 34 (28.81%) | |
| DISCOVERY MR750w | 37 (24.50%) | 80 (47.34%) | 84 (71.19%) | |
| PIRADS | | | | <0.001 |
| 0-2 | 9 (5.96%) | 6 (3.55%) | 1 (0.85%) | |
| 3 | 16 (10.60%) | 6 (3.55%) | 1 (0.85%) | |
| 4 | 15 (9.93%) | 47 (27.81%) | 10 (8.47%) | |
| 5 | 10 (6.62%) | 35 (20.71%) | 77 (65.25%) | |
| Radiographic T-stage | | | | <0.001 |
| T1 | 9/50 (18.00%) | 6/94 (6.38%) | 1/89 (1.12%) | |
| T2 | 34/50 (68.00%) | 66/94 (70.21%) | 32/89 (35.96%) | |
| T3a | 6/50 (12.00%) | 20/94 (21.28%) | 39/89 (43.82%) | |
| T3b | 1/50 (2.00%) | 2/94 (2.13%) | 17/89 (19.10%) | |

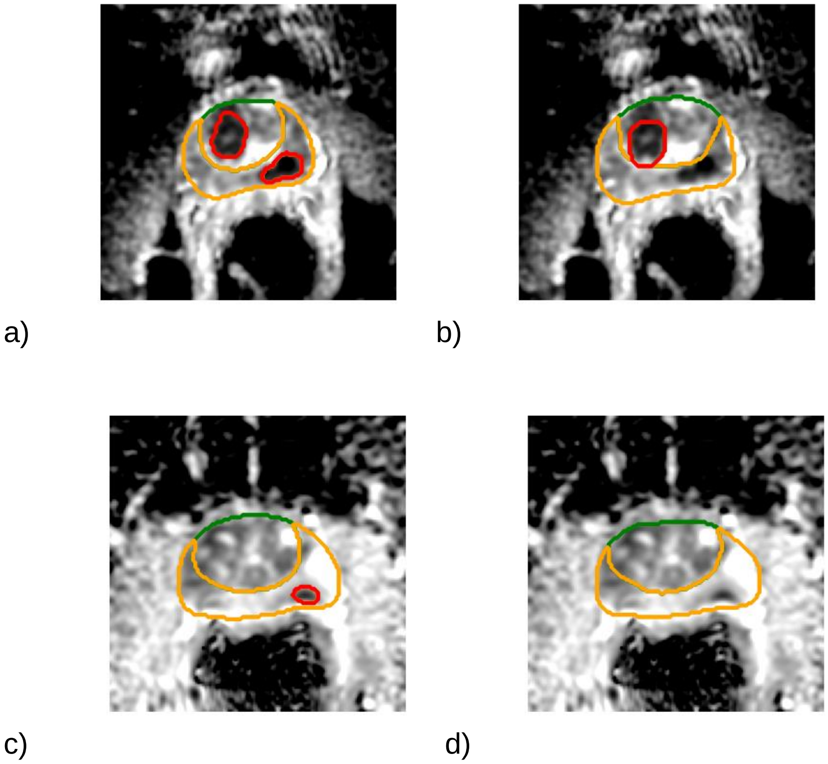


Fig S1: Example of AI (a, c) and reference (b, d) DILs. The green, orange, and red contours correspond to the TZ, PZ, and DIL, respectively. In a), the AI algorithm detected a R TZ-ant PI-RADS 5 lesion, as well as a L apical lesion, which was not present in the reference image (b). However, GS 3+4 disease was present in the L apex on sextant biopsy. In c) the AI algorithm delineated a DIL in the left apex, which was not present in the reference image (d), and could be attributed to an artifact from the endorectal coil.

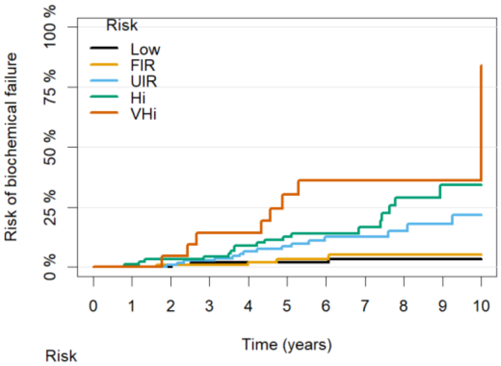


| Risk | | | | | | | | | | | |
|---|---|---|---|---|---|---|---|---|---|---|---|
| Low: | 107 | 104 | 99 | 96 | 91 | 80 | 70 | 63 | 55 | 43 | 14 |
| FIR: | 105 | 103 | 99 | 96 | 88 | 69 | 51 | 42 | 38 | 21 | 6 |
| UIR: | 114 | 114 | 109 | 103 | 94 | 71 | 52 | 41 | 28 | 23 | 7 |
| Hi: | 91 | 88 | 85 | 82 | 77 | 61 | 40 | 29 | 18 | 11 | 5 |
| VHi: | 21 | 21 | 20 | 18 | 17 | 12 | 8 | 5 | 3 | 1 | 0 |

a)

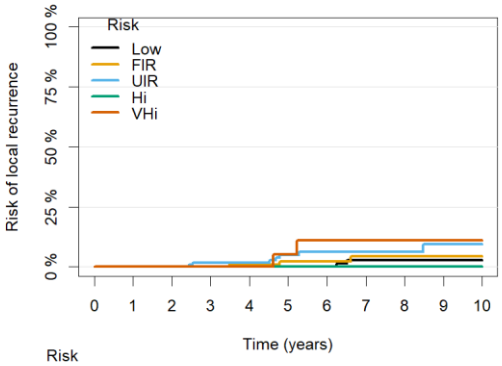


| Risk | | | | | | | | | | | |
|---|---|---|---|---|---|---|---|---|---|---|---|
| Low: | 107 | 104 | 99 | 98 | 93 | 82 | 72 | 64 | 56 | 44 | 14 |
| FIR: | 105 | 103 | 100 | 97 | 89 | 69 | 51 | 42 | 38 | 21 | 6 |
| UIR: | 114 | 114 | 110 | 104 | 99 | 74 | 54 | 41 | 28 | 23 | 9 |
| Hi: | 91 | 89 | 88 | 85 | 84 | 69 | 48 | 36 | 25 | 15 | 7 |
| VHi: | 21 | 21 | 21 | 21 | 20 | 16 | 11 | 7 | 4 | 1 | 0 |

b)

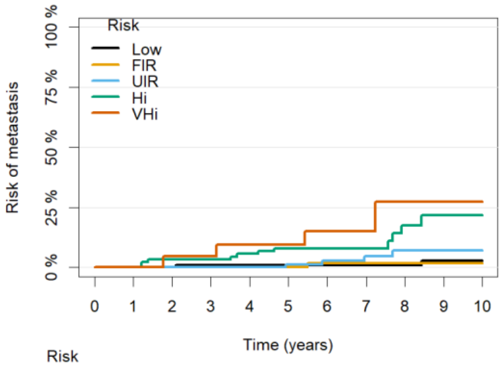


| Risk | | | | | | | | | | | |
|---|---|---|---|---|---|---|---|---|---|---|---|
| Low: | 107 | 104 | 99 | 97 | 92 | 81 | 71 | 65 | 57 | 44 | 14 |
| FIR: | 105 | 103 | 100 | 97 | 90 | 71 | 52 | 44 | 38 | 21 | 6 |
| UIR: | 114 | 114 | 110 | 106 | 101 | 78 | 56 | 42 | 28 | 24 | 8 |
| Hi: | 91 | 89 | 85 | 83 | 80 | 65 | 45 | 33 | 21 | 13 | 6 |
| VHi: | 21 | 21 | 20 | 20 | 18 | 16 | 10 | 7 | 4 | 2 | 1 |

c)

Fig. S2: Association of volume with expanded NCCN risk category (NCCN+) for a) biochemical failure, b) local failure, and c) metastasis.
